# Heterogeneous epigenetic variation converges on splicing dysregulation in opioid addiction

**DOI:** 10.64898/2025.12.20.25342745

**Authors:** Fatir Qureshi, Chesna Apere, Chidera Okeke, Bibi S. Kassim, Marina Iskhakova, Richard Sallari, Maharshi Chakraborty, Laura Morgan, Zia Barnard, Xochitl Luna, Megan Madden, Pavanna G. Rotti, An Hoang, Hannah K. Ramcharan, Bryan Quach, Caryn Willis, Brion S. Maher, Deborah Mash, Peter C. Scacheri, Eric O. Johnson, Schahram Akbarian, Olivia Corradin

**Affiliations:** Whitehead Institute for Biomedical Research; Department of Biology, MIT; Department of Psychiatry Friedman Brain Institute, Icahn School of Medicine at Mount Sinai; Department of Neuroscience, Friedman Brain Institute, Icahn School of Medicine at Mount Sinai; Axiotl Inc, Cleveland, OH, USA; Department of Brain and Cognitive Sciences, MIT; RTI International, Research Triangle Park; Bloomberg School for Public Health, Johns Hopkins University; Univeristy of Miami, Miami, FL USA; Department of Genetics and Genome Sciences, Case Western Reserve University, Cleveland, Ohio

## Abstract

Disease heterogeneity presents a major challenge for genetic and epigenetic dissection of complex traits. Neuropsychiatric traits, such as opioid use disorder (OUD), arise from diverse genetic and environmental factors that uniquely combine in individuals, complicating efforts to identify causal genes and enhancers. We identified epigenetic variation linked to opioid overdose by profiling H3K27ac in the nucleus accumbens (NAc) of 91 cases and controls. While standard approaches identified only 156 loci with differential acetylation, machine learning models built on combinations of putative regulatory regions distinguished cases from controls at high accuracy (>0.95) and were significantly enriched for OUD genetic risk. To leverage disease heterogeneity, we defined individual-specific variation in H3K27ac, termed Variant Enhancer Loci (VEL). VELs converged on shared target genes at a five-fold higher rate than prior prefrontal cortex studies. RNA splicing was the top-enriched ontology, including the splicing regulator CELF5. CELF5 knockdown in iPSC-derived medium spiny neurons was linked to collagen proteins and extracellular matrix organization. Collectively, this study provides new insights into opioid addiction and a framework for personalized epigenetic analysis of heterogeneous diseases.

## Introduction

Communities nationwide have been devastated by opioid misuse, addiction, and associated overdose deaths. Among Americans aged 18–44, drug overdose is the leading cause of death, with opioid overdose accounting for the majority of these fatalities [1]. Worldwide, opioid overdose accounts for 69% of all drug overdose deaths. Opioid addiction is defined by an interplay of both genetic and environmental risk factors. Opioid use disorder (OUD) is estimated to be 40-60% heritable in twins-based studies, on par with other neuropsychiatric disorders [2–4]. While recent genetics studies have expanded known risk, many of the genetic variants that contribute to this heritability remain unknown [5–8]. Environmental factors such as early life adversity, drug accessibility, and socioeconomic status have also been linked to increased risk of addiction [9]. In addition, comorbidities, including neuropsychiatric disorders such as generalized anxiety disorder, depression, and PTSD have been associated with OUD and opioid overdose [10]. Collectively, this diverse range of genetic and environmental risk factors results in a highly heterogeneous disorder, which presents a challenge to efforts to reveal the underlying etiology of addiction.

Human tissue studies have the power to directly capture the changes in gene regulation that are prominent during the course of disease, regardless of causality. Identified alterations in gene regulation may be the result of chronic opioid exposure, acute overdose, or reflect environmental exposures that predate opioid exposure, such as high stress in early childhood. Underlying genetic variants may also contribute to altered RNA levels, changes in histone modification occupancy or changes in DNA methylation. As RNA and epigenetic mechanisms operate with distinct kinetics, they may reflect different causal mechanisms. Thus, tissue studies of epigenetics and gene expression offer complementary insights into the origins of disease.

Given the diversity of causal mechanisms and high disease heterogeneity in opioid addiction, we developed an approach, termed Variant Enhancer Loci (VEL) mapping, to identify case-specific epigenetic variation [11,12]. This approach aims to capture inter-individual variability in enhancer and promoter activity that distinguishes each disease case from the expectation set by a panel of healthy control samples. Critically, this approach then identifies genomic regions with a high density of individual-specific gene regulatory changes that are linked to the same target gene using chromatin interaction data. This strategy enables identification of gene targets that are frequently dysregulated in disease cases, often via disruption of distinct enhancer elements, but are missed by standard analytic approaches.

Here, we performed H3K27ac ChIP-seq, a marker of active promoters and enhancers, in 91 post-mortem human brain tissue samples collected from the nucleus accumbens of opioid overdose cases and accidental death controls. The nucleus accumbens NAc, a central node in the brain’s reward and motivation circuitry, is critically involved in opioid reinforcement, craving, and relapse [13]. While individual H3K27ac peaks were poor distinguishers of cases and controls, multi-peak models were able to distinguish opioid cases with high accuracy and identified genetic regions enriched for OUD genetic susceptibility. Leveraging VEL mapping, we identified 50 genomic regions with a significant convergence of individual-specific epigenetic variation, including known and novel gene targets linked to opioid addiction, such as the splicing regulator CELF5.

Finally, we identify DNA sequence features that distinguish opioid overdose epigenetic variation using large language models.

## Results

### Identification of global differences in H3K27ac occupancy between cases and controls in NAc

We performed H3K27ac ChIP-seq profiling on post-mortem tissue samples from the nucleus accumbens of individuals who died of opioid overdose and had a history of opioid misuse (opioid cases), as well as a cohort of age- and sex-matched accidental death controls screened negative for substance use disorders (SUDs) (controls) [11,12]. Samples with known comorbid neuropsychiatric disorders were excluded. After quality control (see Methods), genotype cross-verification, and covariate matching, 47 case samples and 44 control samples were retained for analysis (Supplemental Figure 1, Supplemental Table 1). Sequencing was performed to a mean read depth of 80,890,693 reads per sample (s.d. = 36,816,647), with no significant difference in read depth between cases and controls (Welch’s two-sided t-test, P = 0.39). To our knowledge, this dataset represents the most comprehensive H3K27ac ChIP-seq study of the nucleus accumbens.

Across all samples, we detected 655,505 unique H3K27ac peaks and quantified H3K27ac using inherent normalization, an approach that leverages positive and negative control regions to normalize ChIP signal between experiments (Methods). We applied standard linear regression incorporating covariates, including age, postmortem interval, genetic ancestry, and sex, and identified 165 peaks (LR peaks, Supplemental Table 2) with significant differences in H3K27ac between cases and controls (Fig. 1A,B, corrected P-value <0.01). We identified 92 putative target genes of these peaks using a combination of published HiChIP interactions from normal human caudate and distance (genes within 5-kb) [14,15]. Comparison of these genes with published single-nuclei RNA (snRNA-seq) from 10 brain regions revealed medium spiny neuron (MSN) specific gene expression patterns for the majority of target genes (Fig. 1C). Gene ontology assessment using Enrichr revealed a significant association with decreased neurotransmitter release (Fig. 1D) [16].

**Figure 1:**
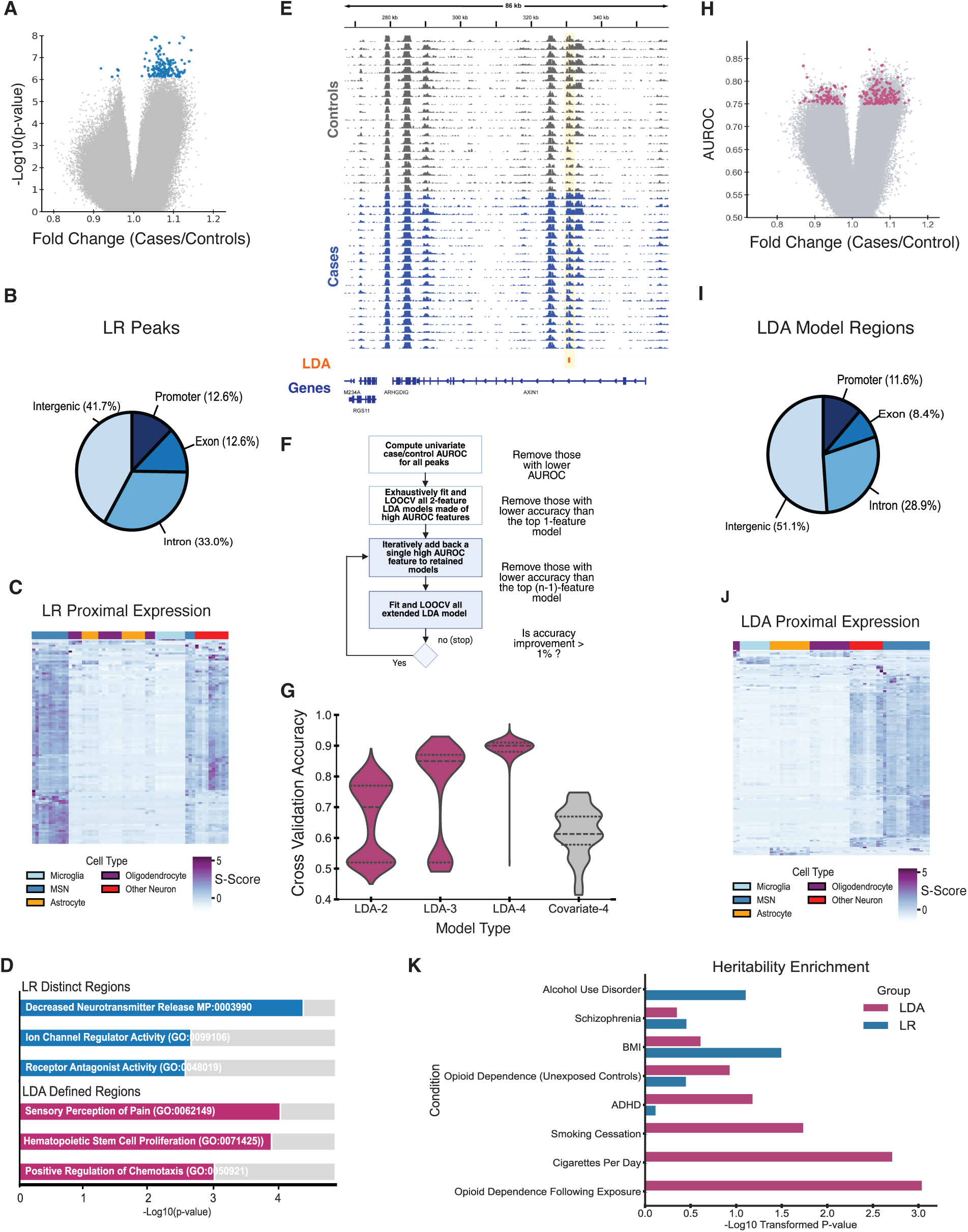
Identification of Global Differences in H3K27ac Occupancy between Cases and Controls in NAc. **(A)** Volcano plot displaying the fold change in the inherent normalized H3K27ac signal between opioid overdose cases and controls. Significant regions (Benjamini-Hochberg adjusted *p* < 0.01) are shown in blue (n=165). **(B)** Genomic annotation of differentially acetylated peaks identified by linear regression (n=165). **(C)** Hierarchically clustered gene expression for genes proximal to linear regression peaks in a previously published single-cell RNA-seq from multiple brains [42]. Expression is stratified by general neuronal populations and MSNs, and glial cell types. **(D)** Gene ontology enrichment analysis of genes proximal to regions identified through both the linear regression and LDA approaches. **(E)** Genome browser displaying H3K27ac ChIP-seq for a subset of opioid overdose cases (blue) and controls (gray). Highlighted in yellow is an H3K27ac peak with high single peak (AUROC=0.76). **(F)** Schematic overview of the linear discriminant analysis (LDA) approach used to identify combinations of regions that distinguish cases from controls. Peaks with high AUROC from univariate classification are selected, followed by exhaustive construction of all two-region LDA models. Models that outperform single-region classifiers during leave-one-out cross-validation are used as inputs for constructing higher-order models. **(G)** Violin plot comparing AUROC distributions for LDA models constructed with 2, 3, or 4 regions, alongside models using only covariates. **(H)** Volcano plot showing inherent normalized H3K27ac fold changes alongside LDA model performance. Regions included in the top-performing 4-region LDA models are highlighted in magenta (n=194). **(I)** Genomic annotation pie chart for regions included in the top 4-feature LDA models, showing their genomic context (n=194). **(J)** Hierarchically clustered gene expression for genes proximal to LDA model peaks in a previously published single-cell RNA-seq from multiple brains [42]. Expression is stratified by general neuronal populations and MSNs, and glial cell types. **(K)** LDSC heritability enrichment analysis showing the contribution of LDA- and LR-identified regions to the heritability of relevant neuropsychiatric and addiction-related traits.

We next examined whether models derived from multiple H3K27ac peaks could better distinguish opioid cases and controls than single peak approaches. We utilized a supervised learning method, linear discriminant analysis (LDA) to maximize the separation between class labels (case vs. control) while minimizing variance within each group [17,18]. Given the high dimensionality of the data, we first performed a feature selection step and identified 6,453 individual peaks that achieved a modest predictive accuracy independently (AUROC > 0.75), with an example shown in Figure 1E. Rather than identify a single globally optimal model, we aimed to uncover diverse, high-performing feature subsets that can distinguish cases from controls (Fig. 1F). To do so, we used a “greedy” search approach [19] to iteratively identify minimal peak sets that accurately distinguished opioid cases from controls. All possible two-feature combinations of peaks were evaluated. Models that outperformed the top single peak classifier based on AUROC value were subjected to leave-one-out cross-validation (LOOCV). This resulted in 1,377 models, comprised of a total of 898 unique peaks which served as seeds for models that extended to 3-peaks. The LOOCV was repeated and features of successful 3-peak models were extended to search for 4-peak models (Fig 1G). Expansion to five-feature models provided only marginal improvement in LOOCV accuracy (<1%) and frequently reduced performance, suggesting overfitting would likely occur beyond 4-peak models. Thus, model optimization was terminated at the four-feature level, resulting in 124 4-peak models comprised of 223 unique peaks, (LDA model peaks, Figure 1H,I, Supplemental Table 2) resulting in a final median model performance of 0.975.

Using HiChIP and distance as described above, we identified 194 putative target genes for these regions, including 12 genes that were also associated with LR peaks. The majority of LDA peak gene targets were specifically expressed in neurons, primarily MSNs. We also observe a minority of genes with astrocyte, oligodendrocyte and microglia specific expression patterns (Fig. 1J). Gene ontology analysis revealed LDA peak gene targets were enriched for GO terms sensory perception of pain and positive regulation of chemotaxis (Fig. 1D).

We next evaluated the genetic variation that coincides with LR peaks and LDA model peaks. Using stratified linear disequilibrium score regression (s-LDSR), we calculated heritability enrichment across 96 traits encompassing neuropsychiatric disorders, substance use related phenotypes and control metabolic and autoimmune diseases (Fig 1K, Supplemental Table 3) [7,20–23]. LDA model peaks were significantly enriched for genetic risk for OUD in a GWAS study that used individuals who were exposed to opioids but did not develop OUD as controls [24] (p-value adj < 0.01). OUD genetic studies that define controls without knowledge of prior exposure status did not show significant enrichment. [25,26]. Smoking-related phenotypes, including cigarettes per day and more modestly, smoking cessation were enriched in LDA peaks [27]. No significant heritability enrichment was observed in LR peaks.

To validate the robustness and specificity of the heritability enrichment observed in the LDA identified peak regions, we evaluated whether this result was common in other sets of H3K27ac peaks identified in the NAc. We randomly selected size-matched peak sets from the full set of NAc H3K27ac identified peaks and calculated heritability enrichment for each set. Across 20 independently selected random sets, none exhibited significant OUD heritability enrichment (Supplemental Fig. 1). These results suggest that chromatin regions contributing more than expected to OUD heritability are preferentially incorporated into optimized multivariate models that separate cases from controls.

Taken together, these findings indicate that no single feature, i.e. H3K27ac peaks, can universally distinguish all cases from controls. Instead, combinations of features are needed to successfully distinguish opioid cases. This suggests a high degree of individual biological variation. However, machine learning classifiers like those we leveraged here, aim to identify the minimal set of features that aid in classification rather than to actively identify all disease-relevant biological variation. Thus, we next sought to identify individual case-specific epigenetic variation systematically.

### Individual-specific epigenetic variation converges on splicing regulators

We reasoned that the diverse genetic, environmental and comorbid contributors to opioid addiction may result in a high degree of heterogeneity across opioid cases. Thus, identifying case-specific alterations has the potential to reveal novel disease-relevant alterations. We previously devised an approach to identify individual-specific epigenetic variation [11,12]. Briefly, this approach uses information theory and random permutation analysis to evaluate each individual case and identify H3K27ac peaks that differ significantly from the expectation set by the full panel of controls (Fig. 2A). We term these regions, Variant Enhancer Loci (VELs) and identified 37,652 hypoacetylated regions, termed “lost VELs” and 16,018 hyperacetylated regions, termed “gained VELs” across our opioid cases (Supplemental Table 4). Consistent with our prior study of the DLPFC, the majority of VEL events were specific to individual cases, with 15% identified in multiple cases (Fig. 2B). However, when we then linked VELs to putative target genes using caudate HiChIP data we found that 60% of VEL target genes were common to multiple cases (Fig. 2C). This suggested VEL regions were not randomly distributed across the genome, but rather converged on specific genetic loci and frequently were in 3-dimensional contact with the same promoter. This pattern showed striking consistency with our prior study of the DLPFC that first demonstrated the convergence of individual-specific variation on shared target genes (Fig. 2B,C) [11].

**Figure 2:**
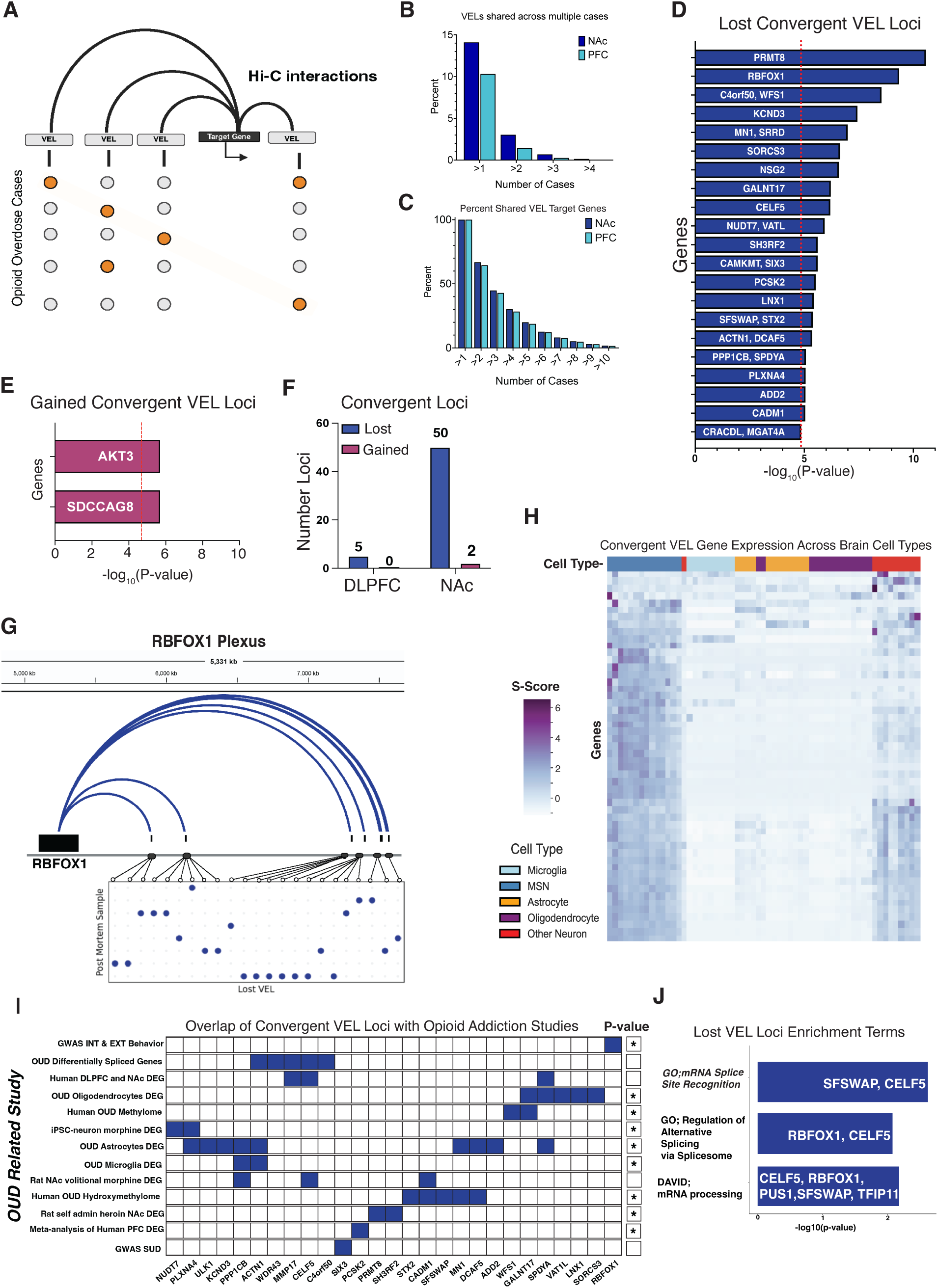
Individual-specific epigenetic variation converges on splicing regulators. **(A)** Schematic illustrating variant enhancer loci (VELs) indicated converging on a shared target gene. Each row represents an opioid overdose and each column represents an H3K27ac peaks. Identified VELs are indicated by orange circles. Arcs represent Hi-C interactions that linked different VELs linked to the same gene. Some VELs are shared across multiple cases and some cases contain multiple VELs displaying both horizontal and vertical convergence. Diagonal convergence is indicated by yellow highlight. **(B)** Percent of VELs found in multiple cases in the Nucleus Accumbens (NAc) cohort and the previously published Dorsolateral Prefrontal Cortex (DLPFC). **(C)** Percent of VEL target genes identified in multiple opioid overdose cases in the two brain regions. Top convergent VEL-associated genes identified through plexus analysis of lost VELs **(D)** and gained VELs **(E)**. The red line indicates the -log(p-value) threshold used for significance in the convergence analysis. **(F)** Comparison of the number of significant lost and gained convergent loci in two different brain regions. **(G)** Hierarchically clustered gene expression for convergent VEL genes in a previously published single-cell RNA-seq from multiple brains [42]. Expression is stratified by general neuronal populations and MSNs, and glial cell types. **(H)** Example convergent VEL plexus for the target gene RBFOX1, showing lost VELs shared across opioid overdose cases and opioid overdose cases with multiple VELs. **(I)** Matrix showing convergent VEL-associated genes (columns) that also appear in other opioid use or addiction studies (rows). Genes that overlap with external studies are highlighted in blue. Genesets with significant overlap with VEL target genes are indicated with an asterisk (GeneWeaver enrichment p-value). **(J)** Gene Ontology analysis of convergent lost VEL gene targets VEL gene targets driving the enrichment are shown on bars.

To identify genomic regions with a significant VEL convergence, we first defined each gene’s regulatory plexus, i.e., the set of all putative regulatory elements linked to the same target promoter, using HiChIP data and H3K27ac peaks (Methods). We then leveraged random permutation analysis to identify plexi with more VEL events than expected at random chance given the number of elements in the plexus. 21 plexi with significant convergence of lost VELs and were identified using permutation analysis (Fig. 2D, E). 10 plexi contained multiple genes resulting in a total of 50 putative target genes. 1 plexus with significant convergence of gained VELs was identified with two putative target genes AKT3 and SDCCAG8. This represents a 5-fold greater degree of convergence in the NAc than in the DLPFC (Fig. 2F). An example plexus is shown in Figure 2G, where 6 opioid cases had at least 2 lost VELs linked to the target gene RBFOX1. We evaluated the expression patterns of convergent VEL target genes using snRNA-seq of human brain tissue and found the majority of genes to be neuronally expressed, specifically in MSNs (Fig. 2H), while a minority of genes including WFS1 and NUDT7 were expressed in glial cells.

Several genes implicated in convergent plexi had previously been linked to substance use disorders in a variety of studies (Fig. 2I). SNPs in proximity to Gained VEL target gene SDCCAG8, a signaling regulator, have been associated with risk-taking behaviors and opioid exposure in both European and African populations [24,28]. The PI3K/AKT pathway is directly activated by opioid signalling, primarily through the gene AKT1. The direct role of AKT3 in opioid-mediated signaling remains unclear. Lost VEL convergent loci included PLXNA4, PPP1CB and SFSWAP. PLXNA4, linked to axon guidance and chemotaxis, has previously been identified as a differentially expressed gene (DEG) in a iPSC-derived neuron morphine exposure study [29]. Protein phosphatase, PPP1CB, has been previously identified in a rat volitional morphine use model in the NAc [30] and SFSWAP was recently identified in as differentially methylated in neuronal profiling of 5-hydroxymethylcytosine (5hmC) in the orbitofrontal cortex (OFC) [31]. Using the geneset analysis platform Geneweaver, we found convergent lost VEL genes were significantly enriched for genesets identified in prior opioid studies including human DNA methylation and gene expression studies of OUD and rat self-administration gene expression studies (Fig. 2I).

Gene ontology analysis of lost convergent VEL loci identified splicing as the top enriched biological process (Fig. 2J), driven by VEL target genes SFSWAP, RBFOX1, and CELF5. CELF5 is highly expressed in the brain compared to other tissues and has previously been identified as a differentially expressed gene in a single nucleus RNA-seq study NAc of rats in a volitional morphine model and in a post-mortem NAc human study of OUD (adjusted p-value <0.05). [30,32]. This suggests that CELF5 and its downstream targets may be a common contributor to the molecular basis of OUD.

### Dysregulation of convergent VEL target gene CELF5 alters splicing in iMSNs

Given the implication of splicing dysregulation in convergent VEL analysis, we sought to further our understanding of the role of CELF5 dysregulation on the function of neurons in the nucleus accumbens. To this end, we generated iPSC-derived medium spiny neurons (iMSNs) using a modified version of a previously published protocol [33]. Immunohistochemistry confirmed the expression of early ventral sub-pallium markers GSX2, SIX3, FOXP1 and ISL1 on day *in vitro* 21 (DIV21), validating fate specification of neural progenitors to the ventral forebrain (Supplemental Fig 3). At later stages of differentiation, expression of transcription factors SIX3 and GSX2 decreased, while ISL1 and FOXP1 expression remained consistent through to DIV40, mimicking the early human developmental stages of MSNs. Striatal projection phenotype was confirmed by DIV40, via the expression of markers such as DARPP-32 and CTIP2 (Fig. 3A and Supplemental Fig. 3).

**Figure 3:**
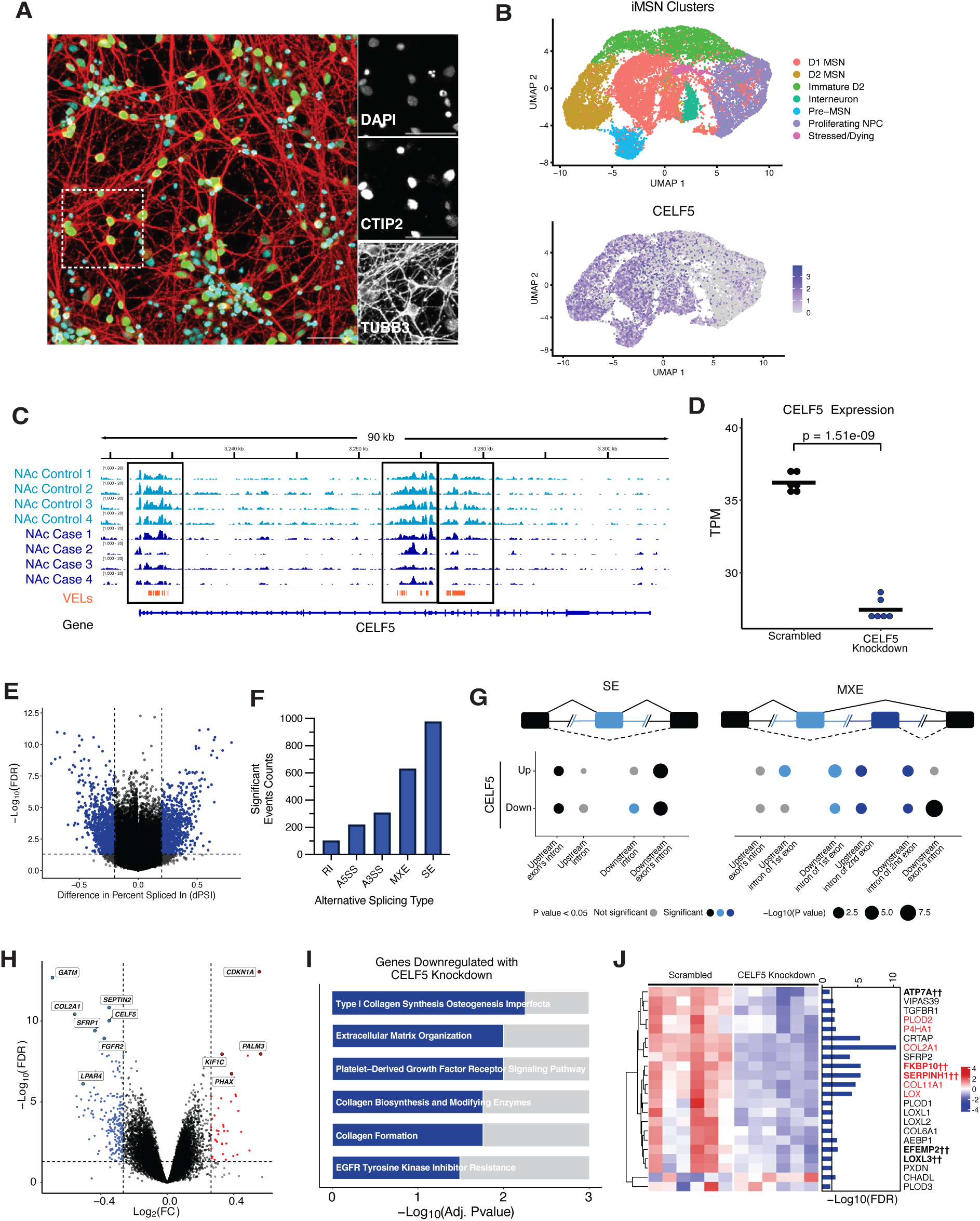
Dysregulation of VEL target gene CELF5 alters splicing in iMSNs. **(A)** Immunofluorescence staining of human iMSNs on day *in-vitro* 40 were stained for canonical MSN marker *BCL11B* (CTIP2, green) and neuron-specific marker *TUBB3* (red) and nuclei counterstained by DAPI (blue). Images were acquired using a ZEISS LSM 980 confocal microscope with a 20× objective and scale bar = 50 µm. **(B)** (Top) Single-cell RNA-seq of DIV39 iMSNs. UMAP of 17,161 cells iMSNs. 7 clusters with predicted cell types are color-coded and identified by expression of canonical cell type-specific markers. (Bottom) Feature plot showing expression of CELF5 across UMAP clusters. **(C)** Genome browser of the CELF5 locus. VEL regions are indicated by orange boxes. 4 representative opioid overdose cases and controls are shown. **(D)** Gene expression (TPM) of CELF5 in CELF5 siRNA and scrambled controls across 6 replicates is shown (P-value 1.5E-9, T-test). **(E)** Volcano plot showing differentially spliced exons after CELF5 knockdown. Significant splicing events are shown in blue (Benjamini-Hochberg adjusted P-value threshold =0.05 and dPSI effect size threshold = 20%). **(F)** Counts of significant differential splicing events per event type. RI = retained intron, A5SS = alternative 5’ splice site of intron, A3SS = alternative 3’ splice site of intron, MXE = mutually exclusive exons, and SE = skipped exon. **(G)** Enrichment of the CELF5 motif at regions flanking the target exon(s) for the top splicing events (SE and MXE). Enrichment for the motif in 250 bp windows of the intron fragments proximal to the target exon (light blue and dark blue) and 250 bp of the intron fragment proximal to the flanking exons (black) are shown. Gray circles = non-significant enrichment; black, light blue, and dark blue circles = significant enrichment. **(H)** Volcano plot showing differentially expressed genes (DEGs) after CELF5 knockdown. Upregulated DEGs are shown in red while downregulated DEGs are shown in blue (Benjamini-Hochberg adjusted P-value < 0.05 and fold-change > 1.2) **(I)** Gene ontology analysis of CELF5 knockdown downregulated DEGs. **(J)** Z-scored expression of collagen-related genes with marginal significance (FDR < 0.1) in CELF5 knockdown. DEGs (FDR<0.05 and fold change >1.2) are highlighted in red and †† signifies genes with significant dPSI exons (FDR < 0.05 and |dPSI| > 20%).

We further assessed the composition of cells generated in the differentiation through scRNA-seq (Fig. 3B). We clustered cells using UMAP dimensionality reduction and assigned likely cell type using expression patterns of known marker genes. We found approximately 25% of cells to be assigned to non-MSN clusters including neural progenitor cells, interneurons and pre-MSNs. The remaining 75% expressed MSN marker genes CTIP2 and OPRM1, half of which expressed known markers of D1 MSNs (ISL1) and half D2 (SIX3) (Supplemental Fig. 4). Terminal maturation of iMSNs was confirmed by co-expression of mature neuronal markers such as MAP2 and ENO2 (Supplemental Fig. 4). Robust CELF5 expression was found across D1 and D2 clusters but not in the NPC cluster (Fig. 3B).

Figure 3C shows lost VELs associated with the CELF5 locus. In order to identify both gene expression and splicing variation linked to CELF5 dysregulation, we used siRNA to knockdown CELF5 expression and paired-end RNA-seq. To account for potential heterogeneity in iMSN differentiations we performed CELF5 knockdown in 6 biological replicates. We found correlation r>0.95 for each of the replicate experiments suggesting a similar proportion of cell types are generated for replicate differentiations (Supplemental Fig 4). We found a 1.3-fold reduction in CELF5 expression across replicates (Fig. 3D). Differential splicing events were evaluated using rMATS [34–37] (Supplemental Table 5). Consistent with other RNA binding protein studies, modest downregulation led to broad impacts on splicing. In total, 2,245 exons with significant differences in splicing were identified with a minimum of 20% difference in percent spliced in (dPSI) (Fig 3E). The majority of dPSI events were skipped exons and mutually exclusive exon use events (Fig 3F). We evaluated the presence of a CELF5 motif surrounding dPSI exons and identified significant enrichment for the CELF5 motif in the introns flanking the target exons for both skipped and mutually exclusive splicing events (Fig 3G). Interestingly, dPSI events were also enriched for RBFOX1 motifs, an additional splicing regulator identified by convergent lost VEL analysis (Supplemental Fig 4).

We detected 258 DEGs with CELF5 knockdown (Fig. 3H, Supplemental Table 5). These genes were significantly enriched for collagen and extracellular matrix (ECM) associated proteins (Fig 3I,J). Prior studies of opioid use disorder including DEGs analysis in the NAc of an independent OUD cohort have implicated ECM associated proteins in opioid addiction [32,38]. We observed modest downregulation of additional ECM genes, including 20 collagen genes (GO ontology:0030199) at FDR<0.07), 5 of which are differential spliced (|dPSI| > 10%). This represents 43% of collagen genes that are expressed in iMSNs (Fig 3J). Collectively, these results suggest that CELF5 plays a critical role in influencing splicing events and ultimately regulation of collagen proteins genes linked to synapse function and cellular communication in MSNs.

### Identification DNA sequence features that distinguish convergent VEL loci

We next sought to determine whether the differences in histone occupancy that we observe at VELs were driven by any shared transcription regulators. To test this, we examined whether DNA sequence features could differentiate lost VELs from other genomic regions. We leveraged DNABERT-2, a large language model (LLM) pretrained on six human reference genomes [39] to compare 150-bp DNA sequences underlying VEL and non-VEL regions identified in the NAc and DLPFC. We randomly sampled training sets of 10,000 VELs and 10,000 non-VELs sequences for each of 25 independent runs and fine-tuned DNABERT-2 to classify VEL from non-VEL regions (Fig. 4A). Models trained to identify DLPFC VELs had minimal success with mean classification accuracy of 51% (s.d. 2%). This is similar to the classification accuracy for models trained on randomly shuffled labels (∼50% accuracy) (Fig 4B). In contrast, models trained to identify NAc VELs achieved a mean classification accuracy of 65% (s.d. 4%). When performance was evaluated on the set of convergent VELs, the mean classification accuracy increased to 71% (s.d. 6%) (Fig 4B). As a comparison to the LLM approach, we performed the same analysis using a k-mer based logistic regression strategy. Regression models were able to distinguish NAc VELs from non-VELs with a mean classification accuracy of 63% (s.d. 1%), but did not exhibit an increased capability to predict convergent VELs (Supplemental Figure 5A). While performance on the training data was comparable between regression and transformer-based models, DNABERT-2 models showed higher average accuracy (Fig. 4C).

**Figure 4:**
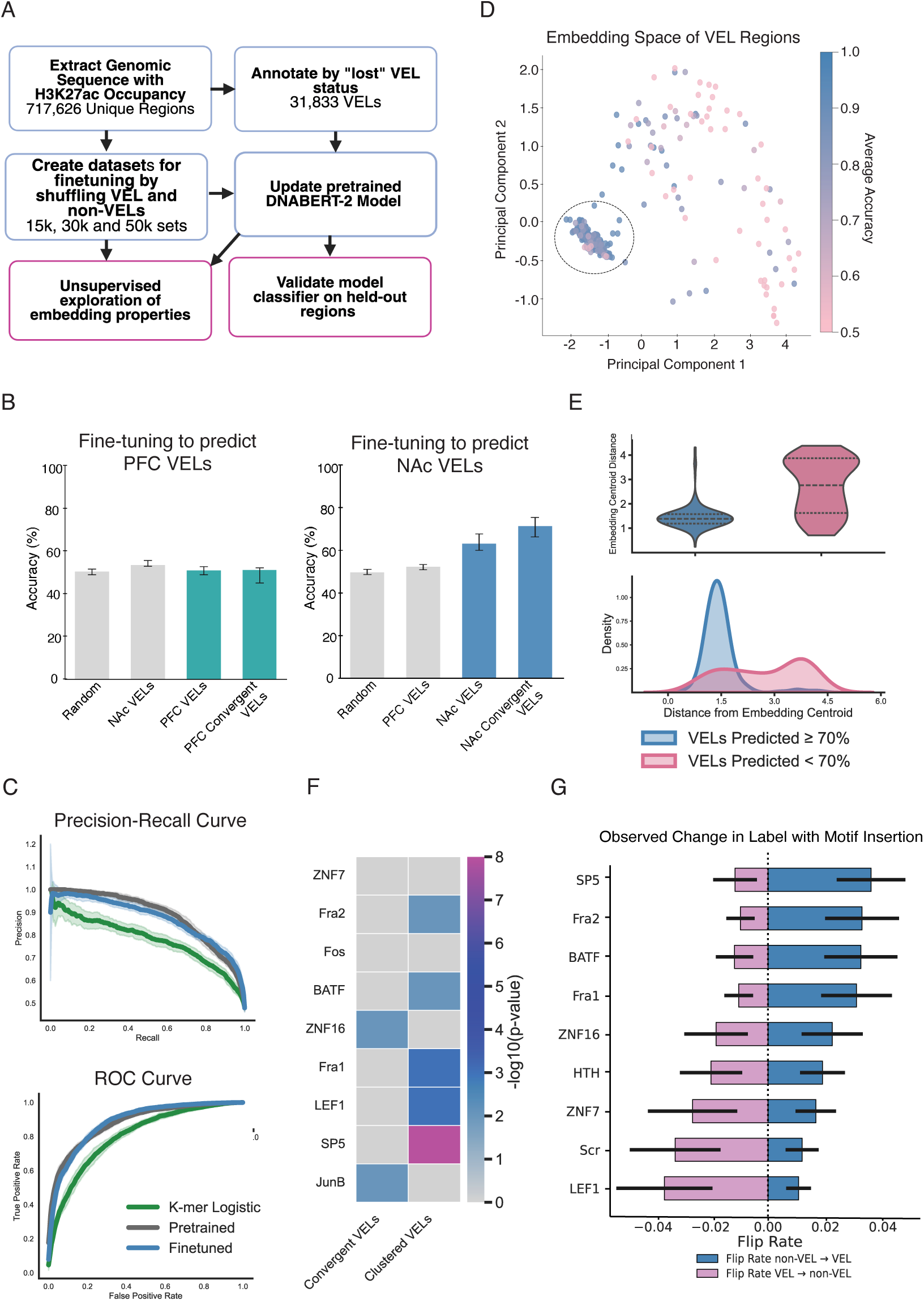
Identification DNA sequence features that distinguish convergent VEL loci. **(A)** Schematic of the workflow to train a foundation model to predict regions of the genome where VELs occur. Genomic regions with significant H3K27ac occupancy were extracted and labeled based on “lost” VEL status across individuals. These were used to fine-tune DNABERT-2 on datasets of various sizes and perform downstream embedding analysis and classification tasks. **(B)** *(Left)* Classification accuracy for models that use DNABERT-2 to distinguish DLPFC VELs from DLPFC non-VELs. Accuracy is shown for control regions (gray) including randomly selected regions and VELs identified in NAc, compared to DLPFC VELs and DLPFC convergent VELs (green). *(Right)* Classification accuracy for models that distinguish NAc VELs from NAc non-VELs. Accuracy is shown for control regions (gray) including randomly selected regions and VELs identified in DLPFC, compared to NAc VELs and NAc convergent VELs (blue). Error bars indicate standard deviation across 20 distinct retrained permutations. **(C)** Performance of DNABERT-2 pre-trained and finetuned classifier methods compared to k-mer logistic regression classifiers on training data. Precision-recall (top) and ROC (bottom) curves are shown. The average AUROC for finetuned (0.88), pretrained (0.88) and logistic regression (0.80) were calculated across 10 independent 20k balanced VEL/non-VEL sequence samples. **(D)** Principal component analysis of DNABERT-2 embeddings from convergent VEL regions, colored by average classification accuracy across bootstraps. The first standard deviation from the centroid of all samples embedding is highlighted by the dashed lines **(E)** *(top)* Violin plot showing the distribution of distances from the embedding centroid, stratified by prediction confidence (VELs predicted <70% vs. ≥70%) (*Bottom*) Density plot showing that more confidently predicted VELs tend to reside closer to the embedding centroid. **(F)** Motif enrichment analysis for all convergent VELs compared to embedding clustered VELs. **(G)** Change in predicted label following insertion of individual motifs into genomic sequences. Bars represent the average probability of change in prediction for VEL to non-VEL (magenta) and non-VEL to VEL (blue) flip rates. Error bars represent standard deviation across 10 iterations.

These results suggest that there may be sequence-based features that distinguish convergent NAc VELs. We hypothesized that the ability of sequence-based models to identify convergent VEL regions may reflect regulation of VELs by a shared transcription factor. We ran standard motif analysis using HOMER on all lost convergent NAc VELs and did not identify significant enrichment for any known TF motifs (corrected q-value >0.3). To investigate whether a subset of convergent VELs are responsible for driving accurate model performance, we performed dimensionality reductions using principal component analysis (PCA) on the embeddings derived from DNABERT-2. The first two PCs together explained on average 69% of the total variance (Fig 4D). We found VELs with high prediction accuracy (>70%, across 25 fine-tuned models) to have lower embedding centroid distances than VELs with low prediction accuracy (<70%) (Fig. 4E). We repeated HOMER analysis on the subset of convergent VELs defined by their embedding distance. When examining only VELs in close proximity in the PCA space (1 s.d of the global centroid), we identified significant enrichment for SP5, LEF1, Fra1/2 and BATF (Fig. 4F, Supplemental Table 6).

To directly assess the impact of these motifs on classification decisions, we employed a SHapley Additive exPlanations (SHAP) inspired motif injection strategy in which transcription factor binding motifs were inserted into sequences annotated as either VELs or non-VELs. We then assessed the “flip rate”, i.e. whether the classifier’s predicted label changed in response to the motif insertion. This revealed SP5, Fra1/2 and BATF to have strong impact on the model classification decision, far exceeding the impact of scramble motif insertion, whereas LEF1 had minimal influence (Fig. 4G). These results implicate SP5, Fra1/2 and BATF as potential drivers of Lost VEL events observed in the NAc.

## Discussion

Traits like opioid overdose and opioid use disorder are highly heterogeneous reflecting diverse genetic and environmental components that collectively impact gene regulation. Genetics, gene expression and epigenetic studies offer complementary strategies to identify genes and pathways critical to disease progression. Here, we applied a comprehensive analytical framework integrating univariate linear models, shallow machine learning, entropy-based approaches, and deep learning. This combined strategy enabled detection of both global patterns and individual-specific alterations that may be overlooked by conventional methods. We demonstrate that machine learning classifiers built on sets of epigenetic features can effectively distinguish opioid cases from controls, despite individual features showing weak or minimal correlation with the phenotype. We further leveraged foundational models to identify underlying DNA sequence patterns associated with epigenetic variation and revealed a new strategy for identifying candidate transcription factor drivers of gene dysregulation. By linking global and case-level variability to overarching biological mechanisms, our approach enhances both the sensitivity and interpretability of complex heterogeneous datasets.

Strikingly, the observation of individual-specific epigenetic variation converging on addiction relevant genes from our prior study of the DLPFC was recapitulated in the NAc. In the DLPFC, convergent VELs localized to genes involved in MAPK, GABAergic, and potassium-channel signaling (ASTN2, KCNMA1, DUSP4, GABBR2), reflecting alterations in neuronal excitability and cortical control. In contrast, in the NAc, convergent VELs are centered on RNA-binding proteins and splicing regulators (CELF5, RBFOX1, SFSWAP), indicating cell-intrinsic remodeling of transcript processing and synaptic architecture. Together these findings suggest region-specific layers of vulnerability — cortical circuits affecting top-down regulation, and striatal circuits affecting structural and transcriptional plasticity. We also observe a dominance of lost VELs compared to gained VELs in both regions suggesting biological continuity across these studies that converges on distinct downstream pathways reflecting diverse regional function. These results highlight that extending future studies to additional brain regions is likely to yield distinct insights into disease biology.

OUD is a multi-stage and cyclical disorder characterized by periods of drug exposure/intoxication, withdrawal, and relapse. Overdose most frequently occurs at the terminal stage of this cycle, typically following relapse. These distinct stages are likely modulated by both overlapping and stage-specific genetic variants, genes and molecular pathways. In aggregate, our study identifies genes implicated in several of these phases. For example, convergent VEL loci demonstrate notable overlap with previous investigations of opioid response. This includes, NUDT7, which encodes a hydrolase that functions to eliminate toxic metabolites from the cell that was found to be differentially expressed in response to morphine in an iPSC-derived neuron model [29]. However, we also observe overlap with model organism studies that distinguish gene expression changes induced by drug exposure from those associated with learned or volitional drug use. This includes CELF5 which is differentially expressed specifically with morphine self-administration rather than acute opioid exposure [30]. Furthermore, our findings reveal an intersection between genetic predisposition to OUD for both convergent VEL and LDA model peaks. Collectively, this suggests the epigenetic changes we identified do not solely reflect changes associated with acute overdose or drug exposure, rather the identified changes reflect multiple distinct layers of the pathogenesis of addiction. Interestingly, this contrasts with gene expression studies that have described DEGs as primarily linked to direct opioid exposure and signaling responses [40,41].

A fundamental challenge in human genetic, transcriptomic, and epigenetic research is the replication of findings across independent studies. Obtaining sufficient sample sizes to robustly identify genes and variants that replicate independently remains a significant barrier. For highly heterogeneous disorders, replication is further complicated by the involvement of diverse causal mechanisms. Consequently, while specific variants, gene regulatory elements, or target genes may not be consistently identified in every study or individual, these disparate molecular alterations can converge to highlight shared molecular drivers of disease biology. The example we highlight here identified CELF5 as a common target of diverse epigenetic dysregulation. Notably, knockdown of CELF5 results in dysregulation of extracellular matrix (ECM) genes, which have previously been reported as differentially expressed in OUD [32]. Convergence may occur at the level of a specific gene, like CELF5 or alternatively, on a downstream gene set such the ECM. Incorporating disease heterogeneity in analytical approaches can enable identification of convergent disease mechanisms at multiple scales. Elucidating such convergent mechanisms is critical for prioritizing targets for the development of future therapeutic or preventive strategies.

## Supplementary Material

Supplemental Figure 1: Global Characterization of Cohorts and Properties of Separatability

Supplemental Figure 2: VEL target gene analysis

Supplemental Figure 3: Immunohistochemistry of key marker genes in iMSNs differentiation

Supplemental Figure 4: Transcriptional analysis of iMSN

Supplemental Figure 5: Logistic Regression Classifier for Predicting VEL Status

Supplemental Table 1: Opioid overdose cohort samples

Supplemental Table 2: Peaks and Target genes identified through linear regression and LDA

Supplemental Table 3: Heritability analysis of LR and LDA model peaks

Supplemental Table 4: Convergent VELs (Variant Enhancer Loci) and Target Genes

Supplemental Table 5: Splicing and transcriptome analysis of CELF5 knockdown in iMSNs

Supplemental Table 6: DNABERT-2 Performance trained on NAc VEL and PFC VELs

## Methods

### ChIP dataset and Opioid Overdose Cohort

#### Opioid overdose cohort and data collection

Post-mortem samples from the nucleus accumbens were obtained from cadaver donors collected by the University of Miami Brain Endowment Bank from multiple ancestry groups. As these samples were anonymized and collected after death, their use does not constitute human subjects research and is therefore exempt from regulation 45 CFR Part 46 (NIH SF424 Part II: Human Subjects).

Opioid overdose cases were defined by stringent criteria applied in characterization in order to account for confounding effects that could influence downstream analysis. The suitability of each sample was determined by forensic pathologists who assessed the cause of death using standard medical and legal procedures. This assessment included review of medical records, police and autopsy reports, and toxicology findings. Urine samples from each individual were subjected to drug screens for common drugs of abuse and alcohol. Positive urine drug screens were confirmed using quantitative assays of both blood and brain tissue. The presence of 6-acetyl morphine (6-AM) was used to confirm acute heroin exposure in individuals who died from opioid overdose.

Samples assigned to the case cohort required documented evidence of opioid use, a positive post-mortem toxicology report for opioids, and a forensic determination that opioid exposure contributed to the cause of death. To further validate each sample and minimize potential confounding, retrospective chart reviews were conducted to gather additional information on opioid use history, participation in methadone or addiction treatment programs, drug-related legal issues, and the presence of drug paraphernalia at the scene of death.

Control NAc samples underwent a conservative selection process to reduce the likelihood of including tissue subjected to acute or chronic opioid exposure. Individuals were excluded from the control group if toxicology reports or medical records indicated any evidence of opioid use. Next-of-kin of individuals included in the control cohort were also contacted to verify no medical history of chronic opioid exposure. Controls were limited to age-matched individuals who died suddenly from causes unrelated to drug use, had no documented history of substance use, and tested negative for major drugs of abuse on post-mortem urine toxicology screens. Subject groups were matched as closely as possible for age, sex, and post-mortem interval (see Supplemental Table 1). Individuals with evidence of prior psychiatric diagnosis, a chronic debilitating illness, or death by suicide were excluded from both cohorts to minimize confounding.

#### Description of genotyping

Genotype data was processed separately for each chromosome (1–22 and X) using a standardized pipeline. Phasing was performed using Eagle v2.4, and genotype imputation was conducted via the Michigan Imputation Server (version 1.5.7) using Minimac4. Separate VCF files were generated for each chromosome and ancestry-defined cohort. For individuals in the European ancestry cohort, imputation was performed using the Haplotype Reference Consortium (HRC) reference panel. For individuals in the African ancestry cohort, the CAAPA reference panels were used.

#### Nucleus accumbens ChIP-seq sample preparation

For each sample, a minimum of 5 x 10^5^ nuclei were extracted from frozen-thawed tissue aliquots by douncing and ultracentriguation, processed by chromatin immunoprecipitation with anti-H3K27ac (Cat# 39133, Active Motif, Carlsbad, CA) and DNA library preparation with KAPA Hyper Prep Kit (KK8502) and BIOO Scientific adapters (Catalog #514102) exactly as described in our detailed step-by-step protocol for postmortem human brain tissue [43].

#### Description of ChIP data

Paired-end reads were processed with Cutadapt v1.9.1 to remove adapter sequences and discard reads shorter than 20 bp [44]. Filtered FASTQs were aligned to the hg19 genome assembly using BWA-MEM v0.7.17-r1188 in paired-end mode with default settings [44,45]. Resulting SAM files were converted to BAM format using SAMtools v1.10 and were subsequently sorted and indexed [46]. Peaks corresponding to H3K27ac occupancy were identified with MACS v2.1.2 using the broad peak setting [46,47]. Signal tracks were generated from final BAM files and normalized to reads per genomic content (RPGC) in 50 bp bins with DeepTools v3.2.0 [46–48]. The bigWig files were visually inspected using the Integrative Genomics Viewer and samples exhibiting abnormal track profiles or low signal-to-noise were eliminated from downstream analysis [49]. Quality control metrics were generated using the ChIPQC Bioconductor package, and libraries with very low mapping rates, RelativeCC enrichment <1, reads in peaks <2%, or fewer than 10,000 peaks were excluded[50]. Peak lists were filtered to remove all peaks overlapping ENCODE blacklisted regions[51].

#### Inherent normalization strategy

Inherent normalization as part of the Axiotl Convergence Platform was used to normalize ChIP-seq signal as previously described [11,52]. This approach uses positive control genomic regions, regions with stable epigenomic activity across reference datasets from the Roadmap 2015 project. Signals within these regions are compared to highly cell type-specific negative control regions (active in only one reference panel cell type). The distance between the average signal across positive and negative controls is used to “inherently” normalize ChIP signal. This method is detailed in Chakraborty et al 2025 and Corradin et al 2022.

#### Linear regression ChIP-seq case/control analysis

Normalized H3K27ac signal intensities were quantified across 773,955 genomic regions in 91 post-mortem nucleus accumbens (NAc) samples and contrasted between case and control cohorts. Covariates included sex, age at death, post-mortem interval (PMI), and ancestry; all covariates were Z-score normalized. Principal components derived via eigen decomposition were identified from genotype data using the *bigsnpr* R package to adjust for population stratification. [53,54]. The number of principal components included as covariates was determined based on a scree plot of explained variance [[53], see Supplemental Figure 1].

Analysis was restricted to individuals that had complete covariate data available. For each genomic region of interest, a linear model was fitted using the inherent normalized H3K27ac signal as the response variable and the covariates as predictors. The resulting residuals, representing covariate-adjusted signal, were then compared between case and control groups using Welch’s t-test (R stats package) [55]. Statistically significant regions were identified using a Benjamini-Hochberg correction, with an adjusted p-value threshold of 0.01.

#### Linear Discriminant Analysis (LDA) classifier based analysis

LDA was used to identify distinct classifier models capable of distinguishing opioid use cases and controls at high accuracy. Initial feature selection screening was performed using the calculated area under the receiver operating characteristic curve (AUROC) between cases and controls for each ChIP region based on normalized H3K27ac signal intensities. AUROC values were computed using the *mltools* R package. Regions with AUROC ≥ 0.75 (n = 6,453) were retained for downstream model development.

LDA models were fit and cross-validated using a standard implementation of the algorithm in MATLAB 2024b. Model construction began with an exhaustive evaluation of all possible two-feature combinations drawn from the retained feature set. Any two-feature model that outperformed the best single-feature classifier based on leave-one-out cross-validation (LOOCV) accuracy, was selected as a seed for further expansion. In subsequent rounds, additional features were added one at a time to each seed model, and all resulting combinations were evaluated using LOOCV. At each step, only models that showed an improvement in LOOCV accuracy compared to their immediate predecessor were retained as seeds for the next iteration. The expansion process continued until adding new features yielded marginal LOOCV accuracy improvements (<1%).

#### Mapping LR and LDA peaks to putative target genes

H3K27ac ChIP-seq peaks were linked to target genes using published interactions identified in H3K27ac HiChIP derived from normal human caudate [14,15]. For peaks not linked by Hi-ChIP, the nearest gene within 2-kb was assigned as the likely target.

#### Human brain single cell gene expression analysis

Previously published single-cell transcriptomic data [42] were used to assess cell type specificity of target genes identified for LDA, linear regression and VEL mapping approaches. To facilitate comparison, gene expression values were standardized using Scikit learn’s StandardScaler with respect to median values across each cluster, allowing for the identification of relative expression differences between cell types.

#### Linkage Disequilibrium Score Regression

Linkage disequilibrium score regression (LDSR) was used to estimate the heritability enrichment of risk taking, neurodegenerative, neuropsychiatric and addiction related traits at genomic regions of interest. Regions of interest including linear regression peaks, LDA model peaks and VEL regions were compiled. SNPs within 2.5kb of H3K27ac peak centers were included. LD scores for the identified SNPs were calculated using European ancestry reference data from HapMap3 and 1000 Genomes Phase 3 (reflecting the ancestry of the GWAS studies), using the approach outlined by Bulik-Sullivan et al. (2015)[56,57].

Summary statistics from relevant GWAS studies were curated from the NHGRI-EBI catalogue. To account for differences in file formatting and ensure harmonization, the Bioconductor based MungeSumstats package was used to process data from each study [58]. The Bulik-Sullivan baseline LDSC model was used to regress GWAS test statistics against the LD scores of the annotated SNPs to obtain heritability estimates. This approach then allowed for the calculation of heritability enrichment by comparing the proportion of total heritability attributable to SNPs within the defined regions to their relative proportion of total heritability captured across the genome for that trait.

#### Gene ontology analysis

Gene ontology and pathway enrichment were evaluated using Enrichr [16,59,60].

#### Variant Enhancer Loci Mapping

VELs were identified using an approach we previously developed [11]. The goal of this technique was to detect regions of H3K27ac enrichment in individual case samples that significantly deviated from cohort-level patterns observed in controls. Each case sample was assessed independently against the full control panel using normalized H3K27ac signal intensities for all chromatin regions of interest.

To quantify the variation of enhancer activity across individuals, we applied a Shannon entropy–based framework. In this context, entropy captures the distribution of signal across samples. For each genomic region, we computed the information content in the case sample and compared it to that of the control cohort to assess whether the case exhibited.

To evaluate whether the observed deviations were greater than expected by chance, we implemented a permutation-based strategy to generate a region-specific null distribution. Case and control samples were first pooled and randomly shuffled. A subset matching the size of the original control cohort was selected to form a permuted control group, from which entropy values were calculated across all regions. One sample was then drawn from the remaining set to serve as a pseudo-case, and its entropy values were compared against the permuted control distribution. This procedure was repeated for 10,000 permutations per region, and later increased to 100,000 to improve the robustness of statistical inference.

For each region, the original case sample’s entropy value was compared to the corresponding null distribution. Regions where the case conveyed significantly more information than expected were classified as “gained” VELs, reflecting potential enhancer activation. Conversely, regions with significantly less information content were classified as “lost” VELs, indicating reduced enhancer activity. This analysis was conducted independently for each case sample across all peak regions, allowing us to identify both gained and lost VELs on a per-sample basis.

#### Convergence analysis

Chromatin interactions between gene promoters and distal non-promoter loci were intersected with a consolidated set of merged, non-overlapping H3K27ac regions derived from ChIP-seq data of opioid overdose cases and matched controls. VELs were classified as either gained or lost in opioid overdose individuals. These VELs were encoded into two separate binary matrices for subsequent analyses.

Convergence analysis was conducted using an entropy-based method implemented in the Axiotl Convergence Platform (convergence.axiotl.com). For each VEL set, permutation testing was performed by comparing the observed convergence statistic from the actual chromatin interaction network (“plexus”) to a null distribution generated through random sampling of plexi from the corresponding binary matrix.

Randomized plexi were matched to the observed data based on promoter activity and connectivity to control for network structure. Global heterogeneity in patient H3K27ac activity was quantified and incorporated into null plexus construction and statistical evaluation to account for interindividual variability.

### CELF5 gene knock-down in IPSC-derived medium spiny neurons

#### Differentiation of human induced pluripotent stem cells into medium spiny neurons

A feeder independent human induced pluripotent (hiPSC) line constitutively expressing dCas9-Zim3-mCherry was differentiated into medium spiny neurons (iMSNs) using a slightly modified variation of a published method [33]. This protocol follows a 40 days in-vitro (DIV) timeline beginning with exposure of hiPSCs to small molecule inhibitors SB431542 and LDN193189, from DIV 0-12 to promote differentiation to the neuroectoderm. On DIV5, the neural progenitors were dissociated using Accutase and re-plated onto Matrigel coated plates with ROCK inhibitor at a density of 300,000 cells/cm^2^ to ensure homogenous population of neural progenitor cells. Cells were then exposed to human recombinant growth factors SHH and DKK1 until DIV 21 to promote fate specification into MSNs. On DIV21, immature neurons were dissociated and replated onto plates coated with Poly-L-Ornithine and Laminin at 10,000 cells/cm^2^ for increased cell recovery and viability. Human recombinant growth factor BDNF was added from DIV 21-40 to promote neuronal maturation and axon elongation. Cells were grown in incubator conditions - 37°C 5% CO_2_ 5%O_2._

#### Immunofluorescence staining of iPSC-derived neurons

On DIV40, cells were fixed with cold 4% paraformaldehyde for 20 minutes (min) at room temperature (RT), followed by three brief washes in phosphate-buffered saline with calcium and magnesium (PBS+/+). Cells were permeabilized in PBS+/+ containing 1% donkey serum and 0.1% Triton X-100 for 7 min at room temperature (RT). Blocking was done in PBS+/+ with 10% donkey serum for 1 hour at RT. Primary antibodies were diluted in PBS+/+ containing 1% donkey serum and incubated overnight at 4°C. The following primary antibodies were used, anti-CTIP2 and anti-TUBB3. The following day, cells were washed three times for 20 minutes each with PBS+/+ and incubated with species-appropriate Alexa Fluor-conjugated secondary antibodies (1:1,000) in PBS+/+ containing 1% donkey serum for 1 h 45 mins at RT in the dark. Nuclei were counterstained with DAPI for 2 min. Fixed cells were imaged in PBS+/+. Images were acquired using a ZEISS LSM 980 confocal microscope with a 20× objective. Image analysis was performed using Fiji (ImageJ, NIH).

#### siRNA mediated knockdown

On DIV37, iMSNs in 6-well plates were exposed to siRNA targeting exons of the *CELF5* gene for 72h. The transfection mixture of Lipofectamine RNAiMAX, siRNA and Opti-MEM I Reduced Serum Medium was prepared according to the manufacturer’s instructions. In vitro spent culture media was aspirated, adherent cells were washed with 1mL phosphate-buffered saline, then 500 µL of Opti-MEM I Reduced Serum Medium was added. A 300 µL of the transfection mixture was added to each well and incubated for 1 hour at 37°C. Cells were then topped up with 3 mL of maturation medium and plates were left undisturbed until the day of RNA collection. On DIV40, spent media was aspirated and cells were washed with 1mL of PBS. Cold Qiazol was added directly to the wells and cells were scraped and transferred to microcentrifuge tubes for storage in −80 °C for subsequent RNA extraction. A scrambled siRNA sequence served as the negative control. Six biological replicates were completed for each condition. RNA was submitted for bulk-RNA sequencing (paired end - 150×150bp, NOVASEQS1, prepared with Watchmaker-mRNA kit).

#### Quantitative Real-Time PCR (qPCR)

Total RNA was extracted from frozen cells stored in Qiazol, using the miRNeasy Mini Kit, according to the manufacturer’s instructions. RNA was eluted in RNase-free water and quantified using a NanoDrop OneC spectrophotometer. Reverse transcription with 150 ng of total RNA was completed as prescribed in the High-Capacity RNA-to-cDNA Kit protocol. Quantitative PCR was performed using TaqMan Gene Expression Assays for *CELF5* and TaqMan Gene Expression Master Mix on the QuantStudio 6 Flex Real-Time PCR System Reactions were set up in 10 µL total volume, containing 1 µL of cDNA, 5 µL of 2× TaqMan Gene Expression Master Mix, 0.5 µL of TaqMan probe, and 3.5 µL of nuclease-free water. There were 6 biological replicates per condition and each sample was run in 3 technical replicates. Thermal cycling conditions were as follows: 50°C for 2 min, 95°C for 10 min, followed by 40 cycles of 95°C for 15 s and 60°C for 1 min. Gene expression was normalized to *ACTB* as the endogenous control. Relative gene expression was calculated using the 2^−ΔΔCt^ method. Samples were compared to control conditions, and data are presented as mean ± standard error mean (s.e.m.) of biological replicates.

#### Single cell RNA sequencing

On DIV39, two wells of iMSNs from a 6-well plate were dissociated using a published protocol [61]. Briefly, stale media was aspirated and adherent cells were washed with 1mL Hank’s Balanced Salt Solution (HBSS). Cells were then dissociated with 1:1 mixture of Accutase and Papain reconstituted with HBSS (1 vial papain + 5 mL HBSS). After 5 minutes incubation at 37°C, cells were collected in 5 mL Wash Buffer containing DMEM/F12 with hepes, GlutaMAX (100X), Bovine Albumin Fraction V (0.1%), ROCK inhibitor (10μM) and DNase/D2 (2mg/mL). Cells were then centrifuged at 150 x g RT for 7 minutes. The supernatant was aspirated and the cell pellet was reconstituted in 1 mL of PBS with 0.04% BSA. This was submitted to the MIT BioMicro Center for 3’ 10X single cell RNA sequencing (sequencing data - 50 + 87 bases pair-end run with 8 + 16 nucleotide indexes).

#### Bulk RNA-seq data processing

FastQC (v0.11.9) [62] was used to examine the quality of the RNA-seq reads. Reads were then trimmed to remove sequencing adapters and low-quality sequences using fastp (v0.23.4) [63]. Trimmed reads were mapped to the human hg38 genome and the GENCODE annotated transcriptome (release V46) with STAR (Spliced Transcripts Alignment to a Reference, v2.7.1a) [64]. featureCounts (v2.0.1)[65] was utilized for read summarization. Lowly expressed genes were filtered out and genes with CPM > 3 in at least six samples were kept. The read counts of the remaining genes were then normalized with the TMM method [66]. edgeR (v3.40.2) [67] was used to perform differential gene expression and differentially expressed genes were called in the quasi-likelihood F-test mode with a cutoff of FDR < 0.05 (Supplemental Table 6).

#### Alternative Splicing Analysis

Differential alternative splicing was assessed across the five major classes of splicing events with rMATS (v4.3.0)[34–37]. The rMATS framework implements a modified generalized linear mixed model to identify differential exon inclusion from bulk RNA-seq data. Junction and exon counts (JCEC) were used as input for quantifying splicing events. To improve effect-size estimation, difference in percent spliced in (dPSI) values generated by rMATS were further processed with the spliceformats package [68], which applies empirical Bayes shrinkage to reduce variance associated with low-coverage or small-sample events. Alternative splicing events were considered significant when exhibiting an absolute corrected inclusion level difference (|dPSI|) > 20% between CELF5 knockdown and scrambled control samples with an adjusted false discovery rate (FDR) < 0.05 (Supplemental Table 6).

Motif enrichment analysis was performed using rMAPS2 (v2.0.0)[69,70] with rMATS output files to identify RNA-binding motifs associated with *CELF5* and *RBFOX1*. Significantly spliced regions were used as target regions for motif enrichment (|dPSI| > 20% and FDR < 0.05), while non-significantly spliced regions (FDR > 0.5; maximum PSI < 85%, indicating that the exon is not constitutively included; and minimum PSI > 15%, indicating that the exon is not constitutively skipped) were used to estimate background binding levels.

Upregulated and downregulated dPSI events were processed separately. For each splicing event, rMAPS2 examined 250 base pairs (bp) of intronic sequence and 50 bp of exonic sequence flanking the splice junctions to assess the positional enrichment of RNA-binding protein motifs. A 50-bp sliding window with a step size of 1 bp was used to count motif occurrences across these regions. The resulting enrichment profiles were filtered, summarized, and visualized in R (Fig 3G, Supplemental Fig 4).

### Identifying DNA sequence features that distinguish VEL regions

#### Fine-tuning DNABERT-2 for VEL classification

DNABERT-2 was used to identify sequence features that may predispose certain regulatory elements to become VELs. The DNABERT-2-117M pretrained model, trained on six complete human genomes to predict bidirectional sequence context, served as the base model used for data exploration [39]. Using the 1000 Genomes Phase 3 as a reference, sequences corresponding to regions with H3K27ac occupancy peaks were extracted. To account for the potential confounding effect of varying tokenization lengths, fixed window sizes extending 75-bp from the peak summits were used. Each region was annotated as either a VEL or non-VEL, and models were constructed using the recommended hyperparameters for DNABERT-2 and its extensions [71]. Required dependencies included the Triton compiler, Keras (Version 2.15) and CUDA-enabled PyTorch for GPU acceleration.

For finetuning, the pretrained model was tasked with correctly classifying a sequence as either having been derived from a VEL or non-VEL region and an equal number of both were used, with 10,000 of each category selected for model finetuning. To ensure robustness, repeated sampling was performed across 50 independent runs. Model performance was also compared to instances where region labels were randomly assigned, rather than reflecting the true outcome. For each model generated, the performance at correctly classifying convergent VELs was evaluated.

Precision and recall for both the original pretrained DNABERT-2-117M and finetuned model was performed using a permutation based strategy.

#### Feature Importance

To assess the contribution of specific sequence motifs to the predictions of the DNABERT-2 model, we employed a SHAP-inspired motif insertion framework. For this process, the model was finetuned independently across ten iterations using different permutations of input training samples to account for stochastic variability in training outcomes. For each iteration, we selected genomic regions, comprising a variable number of VEL and non-VEL regions.

Within each region, candidate motifs were systematically inserted at predefined positions in the input sequences. The effect of motif insertion was quantified by measuring the change in the model’s predicted probability for the target label, calculated as the absolute difference between the prediction for the perturbed sequence and the original sequence. The impact of each motif was summarized by averaging performance across all regions and model iterations. This approach enabled us to identify motifs that most strongly influenced the model’s VEL classification decisions.

#### Logistic Regression

Logistic regression using k-mer features was used to evaluate the robustness of sequence features relevant for VEL prediction, supporting analysis performed using the transformer based DNABERT-2 approach. All analyses using this method were performed using Python (version 3.8) and the Scikit-learn library (version 1.7).Given recent findings that transformer-based models may not consistently outperform less computationally intensive regression-based methods for genomic annotation tasks, this benchmarking provided an interpretable baseline for evaluating sequence-level predictive power [72].

For each genomic sequence, labeled as either VEL or non-VEL, we enumerated all possible k-mers 4 base pairs in length. A feature vector was generated, where each entry indicated the presence or absence of a given k-mer within a sequence, adjusted for the number of instances of each observation. This matrix served as input for training a penalized logistic regression model with L1 regularization (LogisticRegressionCV, scikit-learn) designed to distinguish VEL labeled sequence classes.

The logistic regression model enabled systematic identification of k-mers associated with an increased probability of a sequence being classified as a VEL. To evaluate feature importance, 10 iterations of 15,000 and 20,000 sequence reaches were examined. For each iteration, a 80:20 training/test split cross-validation strategy was used. Motif-associated k-mers were mapped to their corresponding model coefficients, and aggregated across permutations to estimate their mean and variance in predictive weight. For comparative analysis, model performance was benchmarked against the transformer based foundation model DNABERT-2 using the same input sequence and labels.

#### Python and R environments

We used Python v.3.8.10 and R v.4.2.1

## Data Availability

All data produced in the present study are available upon reasonable request to the authors. Processed data is available through https://corradin-chip-nac-opioid.wi.mit.edu/

