## Supplemental Figures for "Heterogeneous epigenetic variation converges on splicing dysregulation in opioid addiction"

**Supplemental Figure 1**

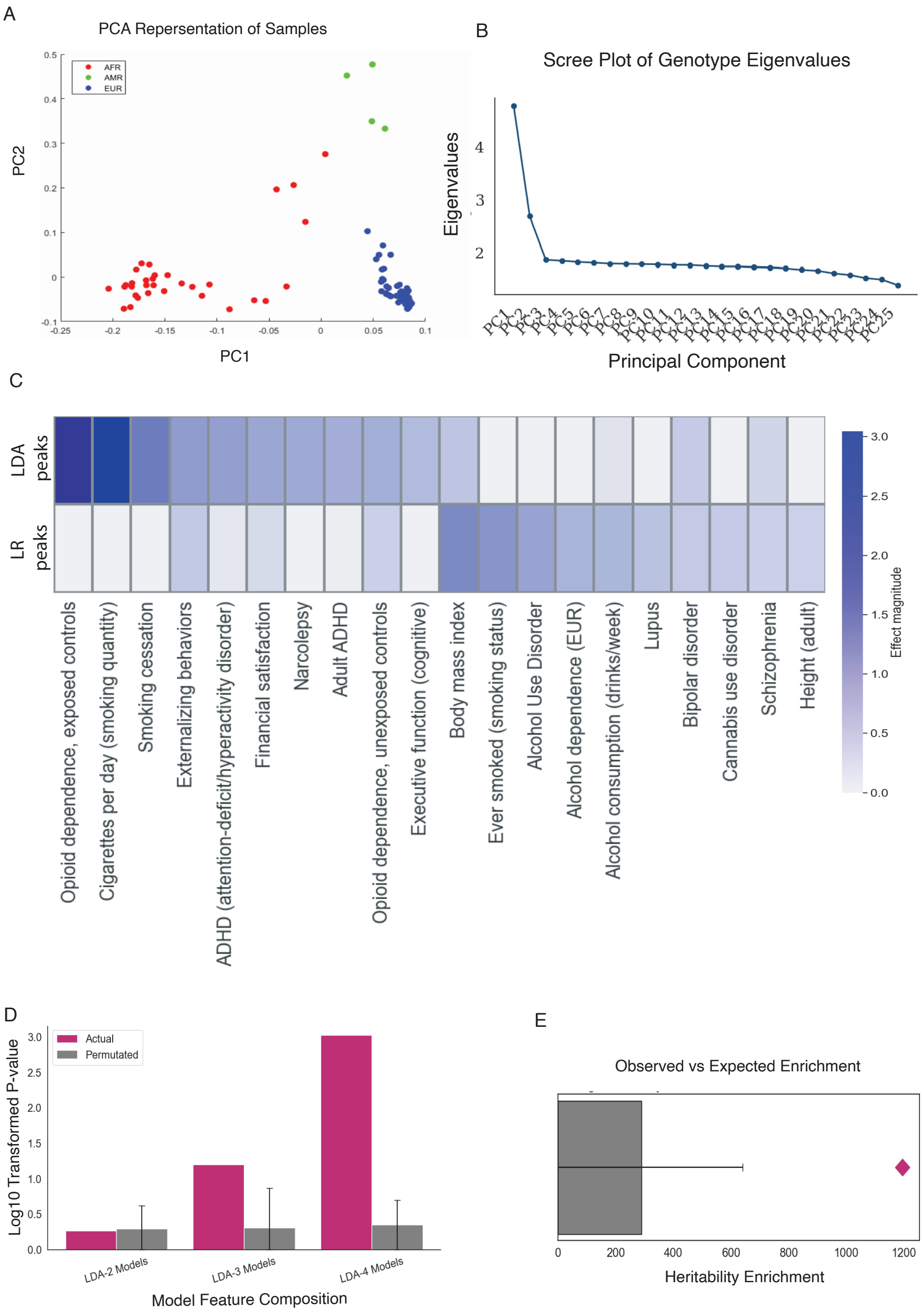


**Supplemental Figure 1. Global Characterization of Cohorts and Properties of Separability**

**(A)** Principal component plot showing the first two PCs derived from genotype data, grouped by cohort described ancestry. **(B)** Scree plot indicating the variance explained by the first ten principal components. **(C)** Extended LDSC heritability enrichment analysis illustrating the enrichment of LDA- and LR-identified regions (see Supplemental Table 3). **(D)** P-values from LDSR-derived opioid use disorder heritability enrichment across top-performing 2-, 3-, and 4-feature LDA models compared to size-matched, randomly permuted regions with known NAc H3K27ac occupancy (n=10). **(E)** Observed versus expected heritability enrichment for the four-feature model compared against the distribution of randomly selected size-matched NAc active peaks.

### Supplemental Figure 2: VEL Target Gene Analysis

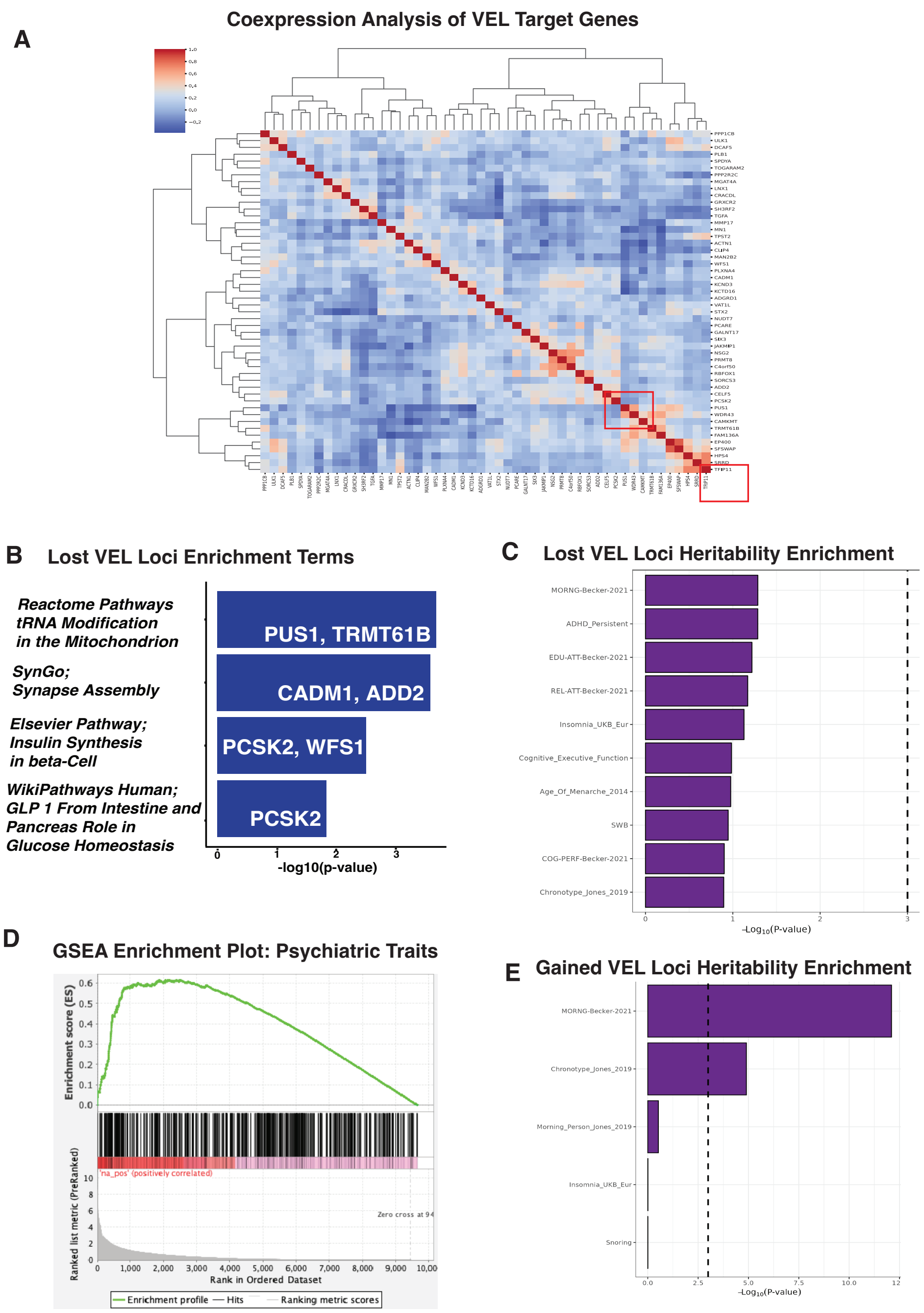


**Supplemental Figure 2: VEL target gene analysis**
**(A)** Co-expression analysis of VEL target genes using the Depmap gene expression dataset. Co-expression of splicing related genes are highlighted in red boxes. **(B)** Extended Gene ontology results for lost VEL target genes. **(C)** Heritability enrichment analysis for Lost VEL and **(D)** Gained VEL loci using s-LDSC. **(E)** Gene set enrichment analysis comparing GWAS SNPs linked to psychiatric traits associated with OUD using VEL genes ranked by convergence p-value.

Supplemental Figure 3: Immunohistochemistry of key marker genes in iMSN differentiation

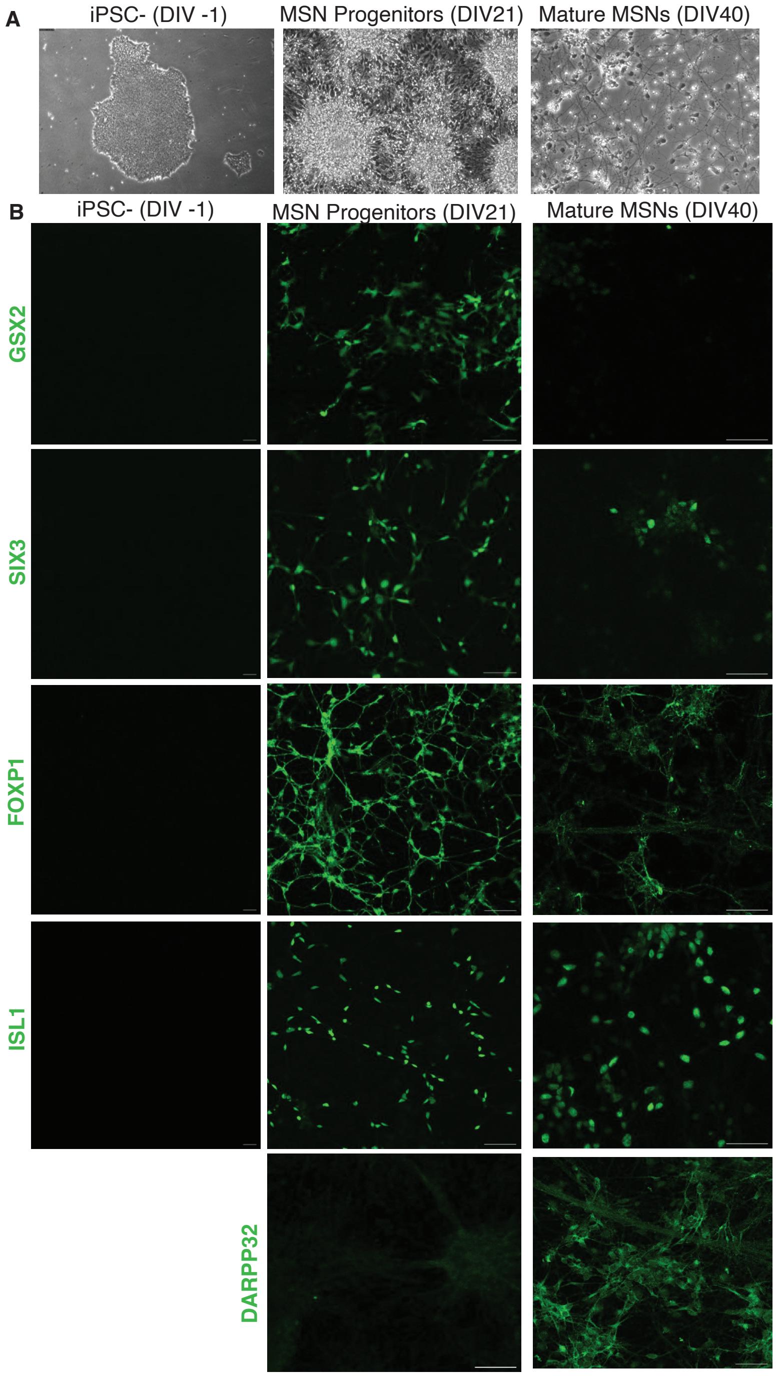


**Supplemental Figure 3: Immunohistochemistry of key marker genes in iMSN differentiation. (A)** Phase contrast images showing morphology changes during iPSC differentiation to iMSNs (DIV1, 21 and 40) are shown. **(B)** Immunohistochemistry for easly ventral sub-pallium markers GSX2, SIX3 and FOXP1 are shown. Later stage iMSN markers ISL1 and DARPP32 are aslo shown. Images were acquired using a ZEISS LSM 980 confocal microscope with a 20× objective and scale bar = 50 μm.

Supplemental Figure 4: Transcriptional analysis of iMSN

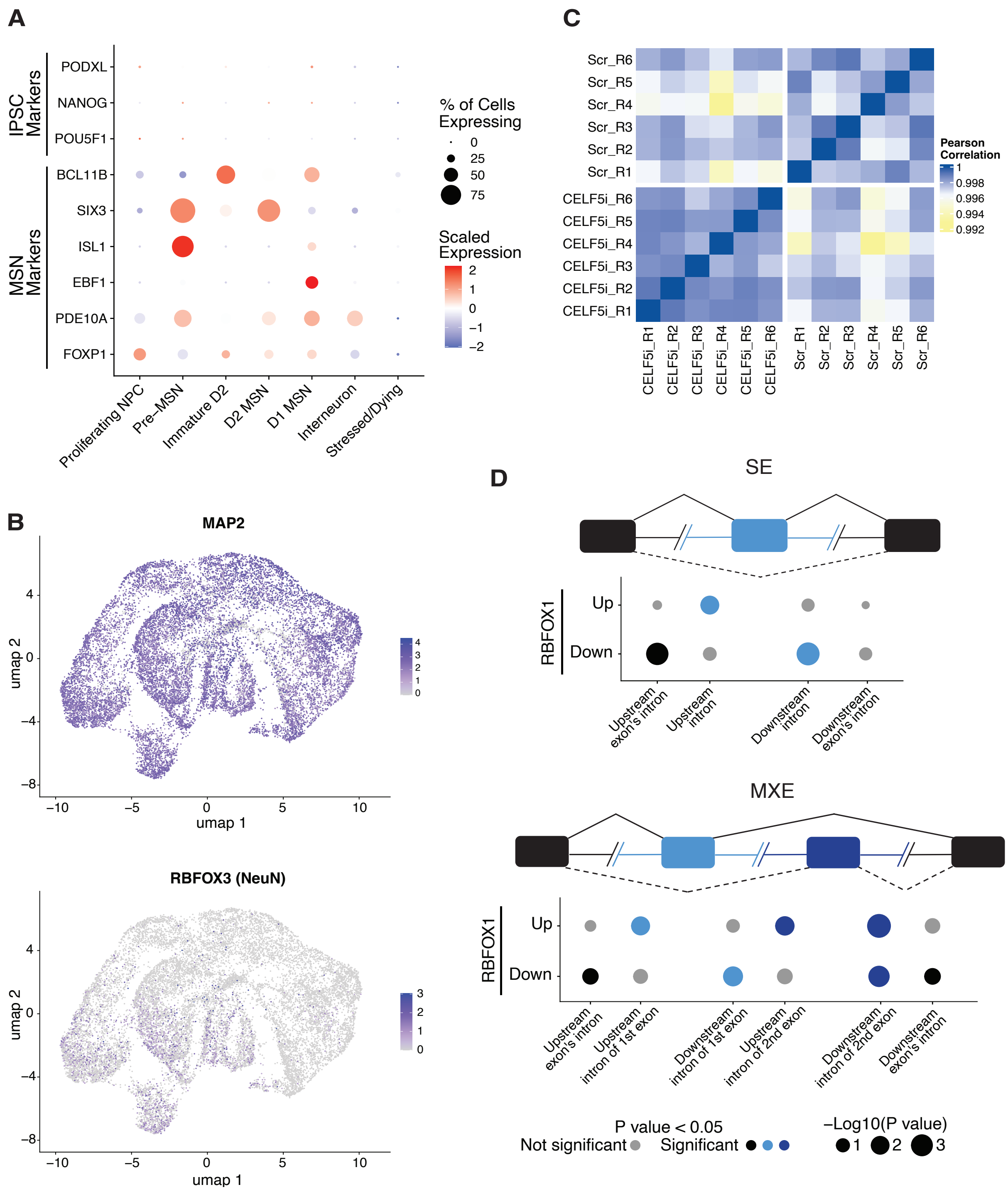


**Supplemental Figure 4: Transcriptional Analysis of iMSNs**

**(A)** Dotplot showing scaled expression of MSN and iPSC marker genes across 7 predicted cell types from iMSN scRNA-seq. **(B)** Feature plot showing expression of neuronal maturation markers (MAP2 and NeuN) across UMAP clusters. **(C)** Heatmap of Pearson's correlation values for the CELF5 knockdown bulk RNA-seq samples. 6 replicates for both conditions (CELF5 knockdown and scrambled control) are shown. **(D)** Enrichment of the RBFOX1 motif at regions flanking the target exon(s) for the top splicing events (SE = skipped exon and MXE = mutually exclusive exons). 250 bp of the intron fragments proximal to the target exon (light blue and dark blue) and 250 bp of the intron fragment proximal to the flanking exons (black) were searched using rMAPS2 and found to be significantly enriched for the RBFOX1 motif. Up and down-regulated dPSI (|dPSI| > 20%) events were computed separately. Gray circles = non-significant enrichment; black, light blue, and dark blue circles = significant enrichment. P-value threshold is 0.05.

Supplemental Figure 5

A

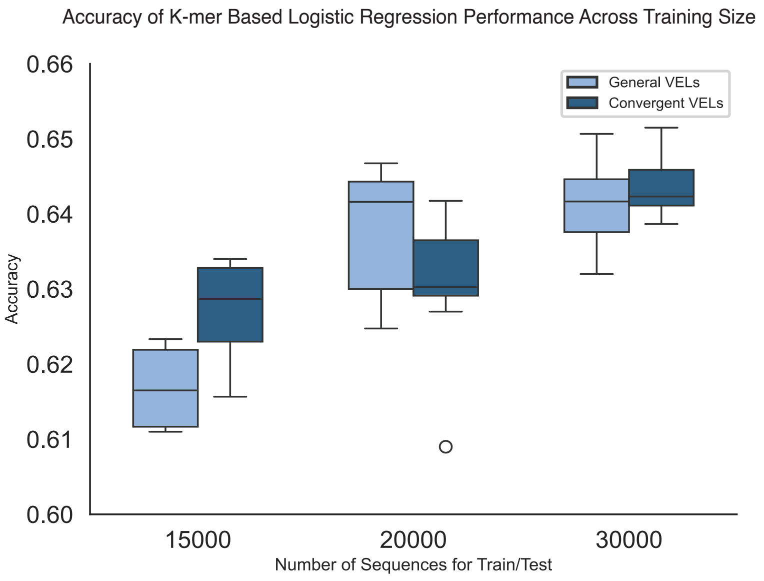


B

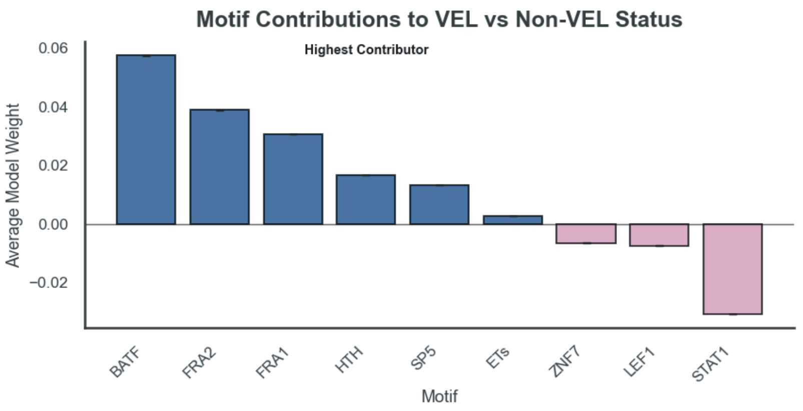


**Supplemental Figure 5: Logistic Regression Classifier for Predicting VEL Status** **(A)** Benchmarking performance of logistic regression using 15k, 30k and 50k sample sets. A K-fold cross-validation strategy was used to evaluate model performance. Performance was validated using both all VELs and convergent VELs specifically **(B)** Model Coefficients assigned to K-mer based weight of motif sequences utilized for label prediction (VEL status). Positive weights correspond to positive label, while negative correspond to alternative.
